# Combining blood transcriptomic signatures improves the prediction of progression to tuberculosis among household contacts in Brazil

**DOI:** 10.1101/2025.09.19.25336212

**Authors:** Sarah Lundell, Vaishnavi Kaipilyawar, W. Evan Johnson, Reynaldo Dietze, Jerrold J. Ellner, Rodrigo Ribeiro-Rodrigues, Padmini Salgame

## Abstract

Tuberculosis remains a major health threat, infecting nearly a third of the world’s population. Of those infected, 5-10% progress from latent infection to active tuberculosis (TB) disease and biomarkers to identify which individuals will progress are needed to allow targeted prophylactic treatment. Several risk biomarkers have been developed to predict progression but have not been tested head-to-head on the same platform. Here, we used the NanoString platform and compared the performance of 15 published gene signatures in predicting progression at baseline in a household contact cohort. Expression of gene signatures was profiled in RNA extracted from whole blood and scored using GSVA and PLAGE. We found that specificity is enhanced by combining signatures and report that the performance of a combined signature that includes a newly derived parsimonious signature through machine learning and a published signature met WHO TPP levels for a triage test. The combined signature had a 90.9% sensitivity and 88% specificity with a PPV of 0.24 and NPV of 1. This combined signature has potential clinical utility in identifying high-risk individuals for targeted prophylaxis to prevent TB morbidity and mortality.

## INTRODUCTION

In individuals exposed to *Mycobacterium tuberculosis* (Mtb), infection leads to a spectrum of potential outcomes, including self-cure, latent infection, subclinical disease, and active TB disease. Of those infected, 5-10% progress to active disease, and biomarkers are needed to identify which individuals will progress, allowing for targeted prophylactic treatment and decreasing the healthcare burden and toxicity associated with treating all latent infections. Current host biomarkers of risk of progression to tuberculosis (TB) disease do not meet WHO target product criteria. A pan-African 16-gene signature (RISK16) was the first biomarker identified in an Adolescent Cohort Study (ACS) that predicted progression within 12 months with 53.7% sensitivity and 82.8% specificity [1]. This biomarker was limited by its accuracy, increasing with decreasing lead time to diagnosis, potentially detecting subclinical disease rather than predicting progression at baseline. Other transcriptomic risk biomarkers are similarly limited by only predicting progression close to the onset of active disease, which may prevent utility in identifying progressors multiple years before the development of active disease [2, 3]. RISK11, derived from RISK16, was found to have limited prediction ability 15 months before progression and only met WHO TPP at a 6-month lead time [4]. RISK11 has also been shown to have a non-specific signal in viral infections [5]. RISK6, a signature derived from the ACS discovery cohort, did not meet the WHO TPP of 12-month lead time to disease progression in validation cohorts [6]. PREDICT29, a transcriptomic signature developed by our group using a dataset from the South African adolescent cohort, predicted progression to active TB in a Brazilian household contact (HHC) cohort at least 5 years before onset with an AUC of 0.911 [7]. This signature remains to be validated in another cohort. In this study, we have validated the performance of PREDICT29 and other published signatures in predicting progression multiple years before the onset of disease in a household contact cohort using the NanoString platform and Gene Set Variation Analysis (GSVA) and Pathway Level Analysis of Gene Expression (PLAGE) modelling. We also evaluated the predictive performance of NANO6, a signature developed to segregate TB and LTBI [8]. The NanoString nCounter system quantifies target gene expression without amplification bias and allows multiple gene sets to be quantified concurrently from the same sample, enabling direct comparison of the performance of multiple signatures. Furthermore, utilizing the same modelling algorithms across signatures allows for direct comparison. GSVA and PLAGE have been shown to effectively quantify expression of gene sets in the case of TB signatures [9]. In this study, we also used machine learning to derive novel risk signatures, which improved specificity when combined with published signatures [8].

## MATERIALS AND METHODS

### Household contact cohort

Participants were recruited in an observational household contact cohort (HHC) in Vittoria, Brazil, as part of the Tuberculosis Research Unit study. Index cases with active tuberculosis were identified in the clinic and their household members were consented and assessed for Interferon Gamma Release Assay (IGRA) reactivity to detect latent infection. Baseline blood samples from latently infected HHCs were collected in PAXgene Blood RNA tubes and stored at -80°C. Additional data were collected, including age, gender, household exposure, and comorbidities. Four years later, the Brazilian national TB registry was checked for development of active TB in the contacts, with microbiological confirmation, after which final classification was made. In our classification, infected HHCs who did not progress to disease are called non-progressors, and those who progressed to disease are progressors, subclassifying those with rapid progression (within 90 days of blood collection) as co-prevalent TB cases. Of the 273 participants in the study, 9 were classified as progressors, 2 as co-prevalent, and 262 were classified as non-progressors. The time to disease onset in the 9 progressors had a median of 1.5 years (range 9 months-4 years). Demographics are summarized in Supplementary Table 1. Time to disease onset and pulmonary vs. extrapulmonary disease data for each subject are available in Supplementary Table 2.

### Study Approval

The study was approved by the Comite de Ética em Pesquisa do Hospital Universitário Cassiano Antonio de Morais, Brazil, and the Institutional Review Boards of Rutgers Biomedical Health Sciences. Written informed consent and assent in Portuguese were obtained from all study participants as per the consent procedure approved by IRBs from all participating institutions.

### RNA extraction and gene expression quantification

PAXgene samples were shipped on dry ice to Rutgers and stored at -80°C, until they were thawed overnight for extraction in batches. RNA was extracted from the PAXgene samples using the Qiagen PAXgene Blood RNA kit according to protocol; briefly, the tubes were centrifuged to pellet the nucleic acids, the pellet was washed and resuspended for protein digestion, cell debris was removed by filtering, RNA was purified by membrane binding and DNAse treatment, eluted and heat-denatured. RNA was quantified and quality was assessed using a Nanodrop spectrophotometer. RNA was hybridized using a custom NanoString mRNA assay encompassing 75 target genes and 6 housekeeping genes to quantify gene expression with the nCounter SPRINT system. The NanoString Gene Expression CodeSet RNA Hybridization Protocol was followed, and sample RNA was diluted with RNAse-free water to 5 μl of 10 ng/μl for a total of 50 ng run per sample. Samples were hybridized for 20h at 65°C before loading on a Nanostring SPRINT cartridge. NanoString is a novel multiplexed platform using digital color-coded mRNA probes that do not require amplification and is more accurate than real-time PCR due to the avoidance of amplification bias [10]. The panel includes the 29 genes of the PREDICT29 signature as well as genes from 14 previously identified signatures, to allow direct comparison of biomarker performance [1, 2, 6, 11, 12] (Supplementary Table 3). nSolver was used to normalize the expression data in a two-step process, with background thresholding on the geometric mean of the negative control counts, and positive control and housekeeping normalization using the geometric means. Downstream analysis was performed in R.

### Signature Analysis

Heatmaps were generated using the pheatmap package, and Uniform Manifold Approximation and Projection (UMAP) was generated using the UMAP package. Signature scores in the panel were calculated using the TB Signature Profiler R application [9] and signature boxplots were generated. Scores were calculated using two different models, GSVA and PLAGE, to compare progressor and non-progressor samples. Performance was assessed via Area Under the Curve and Receiver Operating Curves. Sensitivity, specificity, positive and negative predictive values, and confusion tables were calculated using the pROC package.

### H2o machine learning and derivation of parsimonious signatures

To increase the specificity of the signatures tested in this study in predicting progression and to reduce the number of false positives, machine learning was used to derive new signatures from the data generated here. H2o was used in R to generate new signatures to predict progression [13]. Briefly, the data were split, 55% into a training set and 45% into a test set, with the same proportion of progressors in each set. The H20 “AutoML” function was used to generate models with distinct algorithm types, including Generalized Linear Models (GLM), Gradient Boosting Machine (GBM), and Deep Learning neural networks. For reproducibility, ensemble algorithms were excluded. Seeds were set for both data allocation and modelling for reproducibility. Performance on training data was assessed and then model performance on the hold-out test data was assessed. Some signatures were curated by including top genes that were shared amongst different models, and some were derived from the top genes of a single model. Variable importance gene lists ranking the contributing genes for each model were obtained and a heatmap showing top genes across all models was generated (Supplementary Figure 1). Variable importance data was then used to generate new signatures through several curation methods, including selecting top genes across many models, the top genes from a single model, and combining top genes from distinct algorithm types. A table of the curated signatures and included genes is shown in Supplementary Table 4.

## RESULTS

### Segregation of Progressors from Non-progressors

Unsupervised hierarchical clustering was first performed on the full dataset to determine if progressors would segregate from non-progressors based on expression of the 75 genes comprising 15 published gene signatures (Figure 1). As shown in the heatmap, two major gene expression patterns were observed, with 10 of the 11 progressors clustering together in one group and the remaining one progressor clustering separately in the second group. Several non-progressors with similar gene expression patterns were interspersed among the progressors (Figure 1), suggesting that some signatures may be more nonspecific than others.

**Figure 1:**
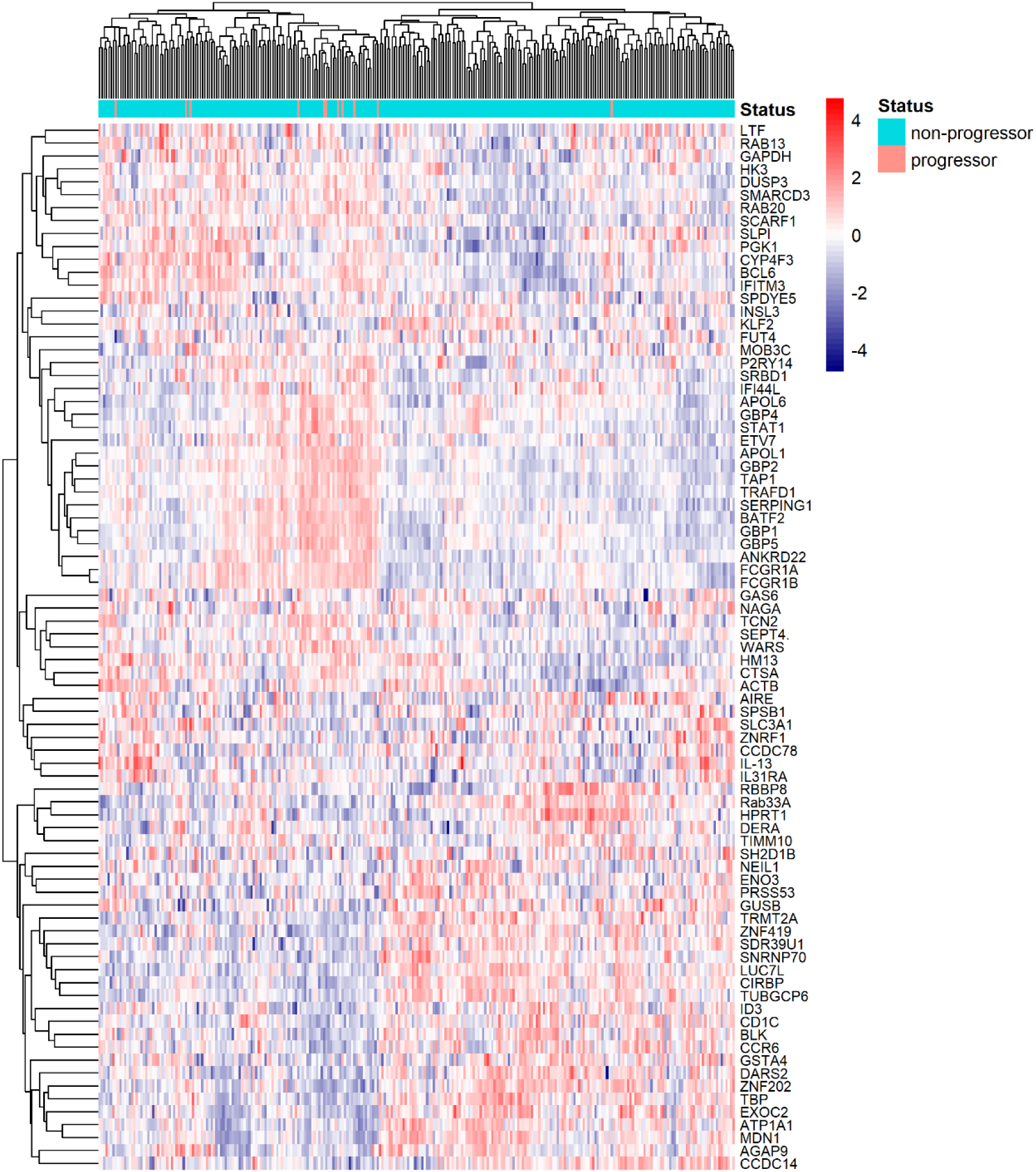
Heatmap of unsupervised clustering of household contacts (n=272) by panel genes.

### Several Published Signatures Predict Progression

We posited that evaluating the performance of each gene signature on the dataset may identify gene signatures with high potential for predictive performance in distinguishing between progressors and non-progressors at baseline. Therefore, we assessed the performance of panel signatures in predicting progression. Signature scores were calculated using the TB Signature Profiler (TBSP) package with the Gene Set Variation Analysis (GSVA) method. GSVA is an unsupervised, non-parametric method that quantifies gene set activity by ranking set genes against all other genes in the data [14]. Signatures that include both upregulated and downregulated genes generally do not perform as well with GSVA, as the oppositely regulated genes can effectively cancel each other out [15]. Thus, PREDICT29, referred to as Leong-RISK29 by the TBSP was subset into Leong-RISK29up and Leong-RISK29down, comprising only upregulated PREDICT29 genes and downregulated genes, respectively. Separating larger signatures into up/down components is a common practice for GSVA. Boxplots of signature scores (Supplementary Fig. 2) showed signature expression differences between progressors and non-progressors, with some signatures better segregating the two groups. Signatures shown to be upregulated in TB, such as RISK11, were more highly expressed in progressors, while those that are downregulated in TB, including NANO6, showed more negative scores in progressors. Area under the curve (AUC) analysis was used to assess performance, and 9 out of the 17 total signatures (including PREDICT29 subsets) showed fair performance, with AUC above 0.7 (Table 1). NANO6 had the highest performance of the set with an AUC of 0.83 (95% confidence interval 0.74-0-0.90). RISK11, RISK16 and Roe3 were also amongst the top performers, with AUCs of 0.83 (0.75-0.89), 0.82 (0.74-0.89) and 0.81 (0.65-0.93), respectively (Table 1). Using GSVA scoring, LeongRISK29-down had better performance than the full LeongRISK-29 signature, with LeongRISK29-up having the lowest AUC of the three (Table 1).

**Table 1.** Ranked table of signature p-values and AUCs using GSVA scoring.

| **Signature** | **P.value** | **-10xLog.P.value** | **LowerAUC** | **AUC** | **UpperAUC** |
| --- | --- | --- | --- | --- | --- |
| Zhao-NANO_6 | 0 | 121.2447 | 0.74 | 0.83 | 0.90 |
| Darboe-RISK11 | 0 | 161.8539 | 0.75 | 0.83 | 0.89 |
| Zak-RISK16 | 0 | 110.5982 | 0.74 | 0.82 | 0.89 |
| Roe-3 | 7.00E-04 | 72.3714 | 0.65 | 0.81 | 0.93 |
| Leong-RISK29down | 2.00E-04 | 85.9127 | 0.69 | 0.78 | 0.87 |
| Kaul3 | 0.0051 | 52.7616 | 0.65 | 0.76 | 0.87 |
| Sambarey-HIV10 | 2.00E-04 | 83.4183 | 0.66 | 0.76 | 0.88 |
| Leong-RISK29 | 0.0022 | 61.3426 | 0.60 | 0.74 | 0.86 |
| Maertzdorf4 | 0.0097 | 46.3905 | 0.56 | 0.74 | 0.85 |
| Sloot-HIV2 | 0.0192 | 39.5201 | 0.56 | 0.66 | 0.77 |
| Leong-RISK29up | 0.0809 | 25.1457 | 0.52 | 0.63 | 0.73 |
| PennNich-RISK6 | 0.096 | 23.4385 | 0.51 | 0.61 | 0.72 |
| Kulkarni-HIV2 | 0.4163 | 8.7631 | 0.51 | 0.59 | 0.77 |
| Suliman-RISK4 | 0.3528 | 10.4178 | 0.51 | 0.58 | 0.73 |
| Francisco-OD2 | 0.393 | 9.3389 | 0.50 | 0.57 | 0.70 |
| Jacobsen3 | 0.3186 | 11.4368 | 0.50 | 0.56 | 0.69 |
| Sweeney-OD3 | 0.8036 | 2.1863 | 0.50 | 0.52 | 0.69 |

Signatures were then scored using PLAGE, a method that quantifies pathway or gene set activity using singular value decomposition. PLAGE is similar to a principal components analysis, and thus can better evaluate bidirectional signatures compared to GSVA [16]. Boxplots of PLAGE scores showed tighter distribution within progressors and within non-progressors (Supplementary Fig. 3). LeongRISK29 had the highest performance of all panel signatures using PLAGE scoring, with AUC of 0.84 (95% CI 0.74-0.93) (Table 2). LeongRISK29, as expected due to its inclusion of upregulated and downregulated genes, showed improved AUC using PLAGE scoring (Table 2) compared to GSVA (0.74) scoring (Table 1). Both LeongRISK29 and LeongRISK29down had higher performance than NANO6 under PLAGE scoring, and these three signatures performed best out of all the panel signatures, with RISK11 and RISK6 also amongst the top performers (Table 2) [6].

**Table 2:** Ranked table of signature p-values and AUCs using PLAGE scoring.

| **Signature** | **P.value** | **-10xLog.P.value.** | **LowerAUC** | **AUC** | **UpperAUC** |
| --- | --- | --- | --- | --- | --- |
| Leong-RISK29 | 0.0119 | 44.3105 | 0.74 | 0.84 | 0.93 |
| Leong-RISK29down | 0.0132 | 43.2628 | 0.76 | 0.84 | 0.93 |
| Zhao-NANO6 | 0.009 | 47.0909 | 0.77 | 0.83 | 0.91 |
| Darboe-RISK11 | 0.0034 | 56.7544 | 0.75 | 0.82 | 0.91 |
| PennNich-RISK6 | 0.0067 | 50.0306 | 0.70 | 0.81 | 0.90 |
| Roe3 | 0.0066 | 50.1559 | 0.68 | 0.81 | 0.89 |
| Jacobsen3 | 0.0066 | 50.2431 | 0.68 | 0.80 | 0.91 |
| Maertzdorf4 | 0.0055 | 51.9505 | 0.70 | 0.80 | 0.92 |
| Zak-RISK16 | 0.0098 | 46.2378 | 0.72 | 0.80 | 0.89 |
| Francisco-OD2 | 0.0231 | 37.6581 | 0.65 | 0.78 | 0.91 |
| Kaul3 | 0.0163 | 41.1549 | 0.65 | 0.77 | 0.91 |
| Sweeney-OD3 | 0.0458 | 30.8269 | 0.61 | 0.75 | 0.85 |
| Suliman-RISK4 | 0.0467 | 30.6492 | 0.65 | 0.74 | 0.85 |
| Sambarey-HIV10 | 0.004 | 55.1248 | 0.61 | 0.74 | 0.85 |
| Sloot-HIV2 | 0.0052 | 52.5905 | 0.61 | 0.73 | 0.83 |
| Leong-RIS29up | 0.0105 | 45.5283 | 0.63 | 0.73 | 0.85 |
| Kulkarni-HIV2 | 0.2466 | 13.998 | 0.50 | 0.60 | 0.76 |

### Thresholding for Sensitivity and Specificity

To assess signature sensitivity and specificity in this cohort, signature score cutoffs were set. The WHO Target product profile for a predictive progression biomarker is balanced for sensitivity and specificity, with a minimum requirement of 75% for both [17]. However, the goal of this test is to identify potential progressors for prophylactic treatment and therefore the risk of a missed case (false negative) significantly outweighs the risk of giving prophylaxis to a non-progressor (false positive), as prophylaxis is already given to all cases of LTBI in low incidence countries such as the United States, and recommended in Brazil [18, 19]. Therefore, a cost-sensitive method was used to select the starting thresholds, prioritizing sensitivity by weighting the relative cost of false negatives versus false positives as the inverse of the estimated Brazil prevalence ratio (5%) [20]. These weights were modified as needed to select thresholds with a sensitivity of at least 80%.

Sensitivity, specificity, positive predictive value (PPV), and negative predictive value (NPV) for these cutoffs were then calculated for the top-performing signatures for both GSVA and PLAGE scoring (Table 3), using the Brazil prevalence rate. NANO6, the top-performing signature under GSVA scoring, had 91% sensitivity and a specificity of 73% (Table 3). Even though NANO6 met WHO TPP levels for sensitivity and was near the specificity level, the PPV was low (0.13), likely due to the low prevalence of progression in Brazil and the non-specificity of host-based signatures [5]. RISK11, the next best GSVA-scored signature by AUC, had a perfect sensitivity of 100% and limited specificity of 65%, with a lower PPV of 0.11 (Table 3). Under PLAGE scoring, the specificities of the top signatures were lower compared to GSVA scoring, except for LeongRISK29 (Table 3). NANO6, the top signature by AUC, had 91% sensitivity and 69% specificity with a PPV of 0.11. RISK11, which is also amongst the top PLAGE signatures by AUC, had 100% sensitivity and a specificity of 62.5%, with slightly lower specificity at the same sensitivity compared to GSVA (Table 3).

**Table 3:** Performance metrics (sensitivity, specificity, PPV, NPV) of selected top-performing signatures at set thresholds using GSVA and PLAGE scoring.

| **GSVA** |  |  |  |  |  |
| --- | --- | --- | --- | --- | --- |
| **Signature** | **Threshold** | **Sensitivity** | **Specificity** | **PPV** | **NPV** |
| Zhao-NANO6 | -0.2912 | 0.91 | 0.73 | 0.13 | 0.99 |
| Darboe-RISK11 | 0.2061 | 1.00 | 0.65 | 0.11 | 1.00 |
| Zak-RISK16 | 0.0740 | 1.00 | 0.64 | 0.10 | 1.00 |
| Roe3 | 0.4101 | 0.82 | 0.72 | 0.11 | 0.99 |
| Leong-RISK29down | -0.2340 | 0.82 | 0.70 | 0.10 | 0.99 |
| Kaul3 | 0.2042 | 0.82 | 0.64 | 0.09 | 0.99 |
| Sambarey-HIV10 | 0.1492 | 0.82 | 0.61 | 0.08 | 0.99 |
| Leong-RISK29 | -0.0396 | 0.91 | 0.58 | 0.08 | 0.99 |
| Maertzdorf4 | 0.1931 | 0.82 | 0.67 | 0.10 | 0.99 |

| **PLAGE** |  |  |  |  |  |
| --- | --- | --- | --- | --- | --- |
| **Signature** | **Threshold** | **Sensitivity** | **Specificity** | **PPV** | **NPV** |
| Darboe-RISK11 | -0.0014 | 1.00 | 0.62 | 0.10 | 1.00 |
| Leong-RISK29 | -0.0033 | 0.91 | 0.66 | 0.10 | 0.99 |
| Zhao-NANO6 | -0.0087 | 0.91 | 0.69 | 0.11 | 0.99 |
| Roe3 | -0.0175 | 0.82 | 0.74 | 0.12 | 0.99 |
| PennNich-RISK6 | -0.0145 | 0.82 | 0.67 | 0.10 | 0.99 |
| Leong-RISK29down | 0.0056 | 0.91 | 0.66 | 0.10 | 0.99 |
| Jacobsen3 | -0.0149 | 0.91 | 0.66 | 0.10 | 0.99 |
| Maertzdorf4 | -0.0253 | 0.73 | 0.75 | 0.11 | 0.98 |

Next, we generated Confusion Matrices to visualize the performance of GSVA and PLAGE scoring. Figure 2 displays the Confusion Matrices in a concise graphical way, showing true positives (TP), true negatives (TN), false positives (FP) and false negatives (FN) for the top two signatures from GSVA (NANO6 and RISK11) and PLAGE (LeongRISK29 and RISK11). The Confusion Matrix for NANO6 (GSVA) demonstrated that at the set threshold, 10 out of 11 progressors would be detected with 70 false positives (Figure 2), leading to the observed low PPV (Table 3). The confusion matrix for RISK11 (GSVA) indicates that all progressors are detected, with 91 false positives resulting from the lower specificity (Figure 2). PLAGE-calculated signatures show that NANO6, at the same sensitivity as calculated in GSVA, has more false positives (82 vs. 70), and similar results were observed for RISK11. LeongRISK29 performed better under PLAGE scoring as expected, due to the higher AUC values, with 90.9% sensitivity in both methods. GSVA-scored NANO6 had the highest PPV of both methods (Table 3). In the context of a triage test for treatment, NANO6 (GSVA) would correctly assign 191 non-progressors out of the 272 household contacts tested in this study, reducing the total number of latent TB to treat by 70%, a more manageable population. Because of the non-specificity, there would be 70 false positives treated alongside the 10 progressors identified (Figure 2).

**Figure 2:**
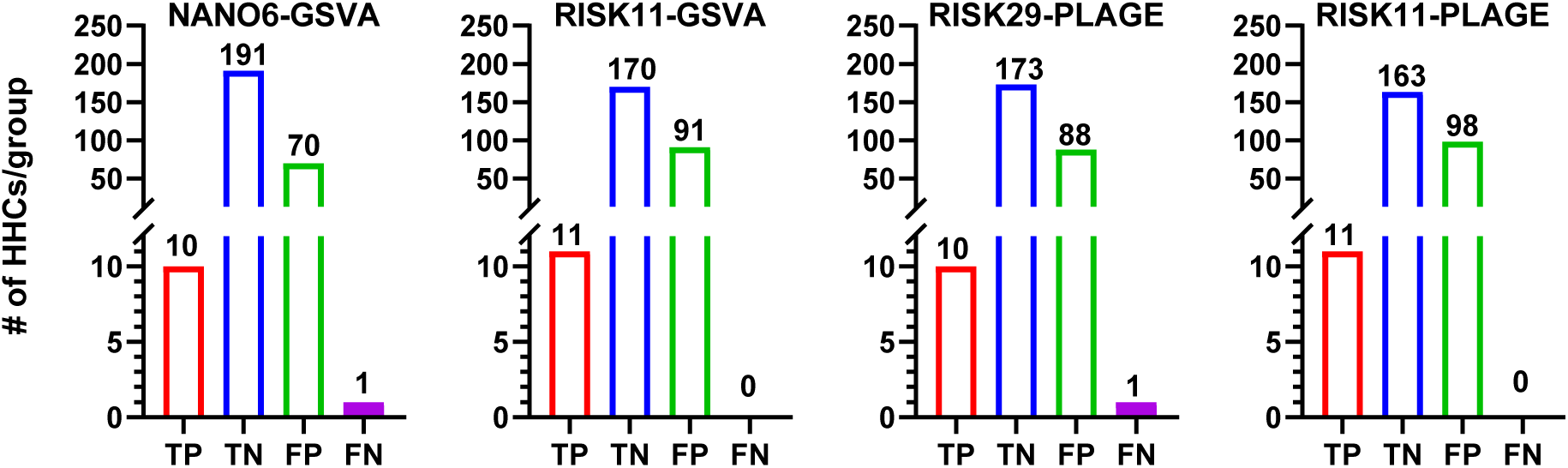
Confusion matrix for selected signatures scored by GSVA and PLAGE with True Positives (TP), False Positives (FP), False Negatives (FN), and True Negatives (TN) presented in a concise graphical way.

#### Improved performance of Machine Learning Derived Signatures compared to Published Signatures in Predicting TB Progression

To increase the specificity of these signatures in predicting progression and reduce the number of false positives, machine learning was used to derive new signatures in this cohort using h2o.ai. A table of the curated signatures and included genes is shown in (Supplementary Table 4). These signatures were then tested in the full dataset using GSVA and PLAGE to directly compare performance to published signatures. GSVA and PLAGE scoring of h2o-derived and published gene signatures on the full dataset demonstrated that several machine learning-derived signatures retained prediction accuracy without their original model algorithms. H2otg showed improved AUC (0.89) compared to the best-performing published signature in GSVA, NANO6 (0.83) (Supplementary Table 5). Although the machine learning produced an improved signature, the AUC suggests that there is inherent non-specificity in the expression of panel genes in this cohort, which prevents complete segregation of the groups. This is consistent with the initial heatmap of the expression data, which showed some non-progressors overlapping with progressors. H2o signature performance was also evaluated using PLAGE. AUC analysis revealed that most h2o signatures performed better than published signatures using PLAGE (Supplementary Table 6), likely due to greater similarity between PLAGE scoring and the original algorithms. Interestingly, the top signature in PLAGE was h2ox1 (AUC 0.89) (Supplementary Table 6), which did not perform well in GSVA (Supplementary Table 5). The top GSVA signature, h2otg, also performed well with an AUC of 0.85, on par with the best published signature under PLAGE scoring (LeongRISK29; AUC 0.84) (Supplementary Table 5).

Signature thresholds were determined as previously discussed, and sensitivity, specificity, PPV, and NPV were calculated for selected top-performing h2o-derived signatures in GSVA and PLAGE scoring and compared to the top published signatures for GSVA (NANO6 and RISK11) and PLAGE (PREDICT29, RISK11 and NANO6). H2otg had a sensitivity of 100%, specificity of 81%, and PPV of 0.18 in GSVA scoring, an improvement over NANO6 (sensitivity 91%, specificity 73%, PPV 0.13) (Table 4). H2ox1, the top performer in PLAGE, had similar performance to h2otg (GSVA), with a sensitivity of 91%, a specificity of 82%, and a PPV of 0.18 (Table 4).

**Table 4:** Performance metrics (sensitivity, specificity, PPV, NPV) of selected top-performing signatures at set thresholds using GSVA scoring and PLAGE scoring.

| **GSVA** |  |  |  |  |  |
| --- | --- | --- | --- | --- | --- |
| **Signature** | **Threshold** | **Sensitivity** | **Specificity** | **PPV** | **NPV** |
| NANO6 | -0.2912 | 0.91 | 0.73 | 0.13 | 0.99 |
| RISK11 | 0.2061 | 1 | 0.65 | 0.11 | 1 |
| h2otg | 0.3535 | 1.00 | 0.81 | 0.18 | 1.00 |

| **PLAGE** |  |  |  |  |  |
| --- | --- | --- | --- | --- | --- |
| **Signature** | **Threshold** | **Sensitivity** | **Specificity** | **PPV** | **NPV** |
| RISK29 | -0.0033 | 0.91 | 0.66 | 0.10 | 0.99 |
| NANO6 | -0.0087 | 0.91 | 0.69 | 0.11 | 0.99 |
| RISK11 | -0.001447 | 1 | 0.62 | 0.10 | 1 |
| h2otg | -0.0111 | 1.00 | 0.70 | 0.12 | 1.00 |
| h2ox1 | -0.0393 | 0.91 | 0.82 | 0.18 | 1.00 |

Confusion matrices were constructed for h2otg and h2ox1. In GSVA, h2otg had improved detection with 0 missed progressors compared to 1 missed progressor with NANO6 (Figure 2), and greater specificity, decreasing the number of false positives from 70 (Figure 2) to 50 (Figure 3). In PLAGE scoring, h2ox1, at the same sensitivity as LeongRISK29, decreased the false positives by nearly a half, from 88 to 46. Although machine learning improved specificity and reduced the number of false positives, there was still non-specificity, with approximately 20% of non-progressors being signature-positive.

**Figure 3:**
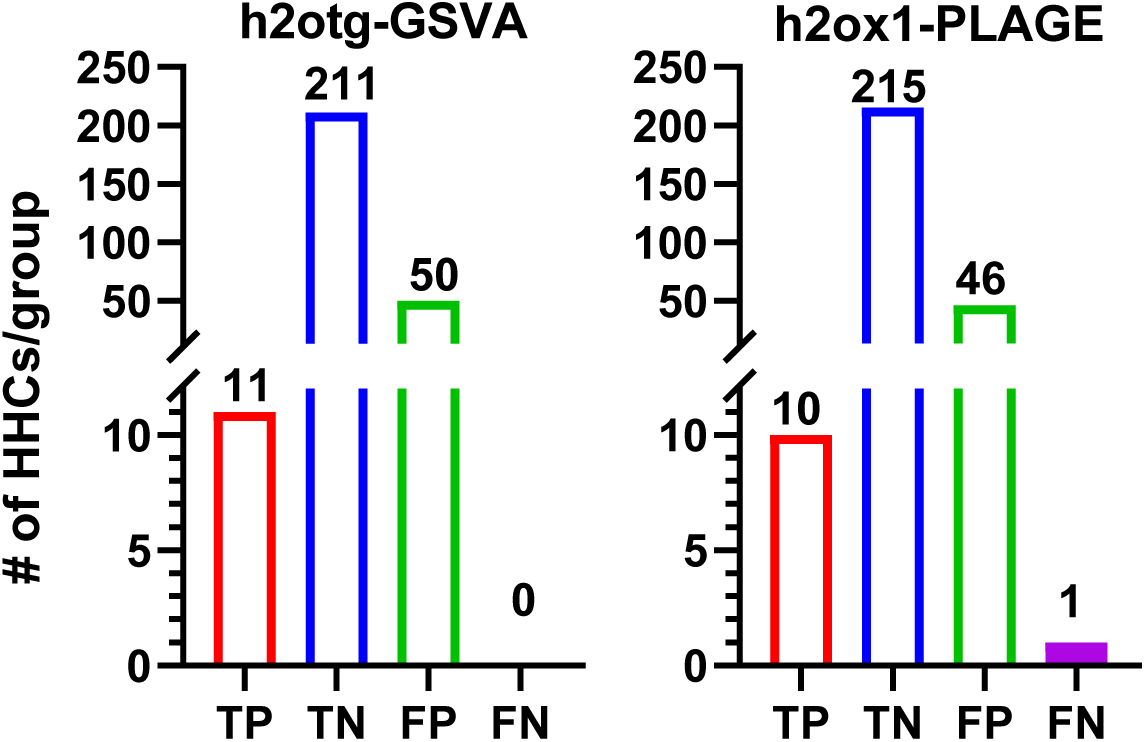
Confusion matrix for Top H2o-derived signature scored by GSVA and PLAGE with True Positives (TP), False Positives (FP), False Negatives (FN), and True Negatives (TN) presented in a concise graphical way.

#### Combination of Top Signatures Improves Specificity

While the standalone signatures markedly reduced the target population for prophylaxis, we hypothesized that we could achieve greater specificity and higher PPVs by requiring positivity on two signatures instead of one. Combining gene signatures has shown improvement in the prediction of Breast Cancer Survival [21], and combining T cell-inflamed gene expression profile with certain myeloid cell-related signatures significantly enhanced the prediction of anti-PD-(L)1 response in non-small cell lung cancer [22]. Hence, by combining information from two gene signatures, one could potentially increase the prediction power.

To further improve the specificity of these signatures and obtain higher PPVs, we tested two-signature combinations, h2otg with NANO6 and h2otg with RISK11 (GSVA), and h2ox1 with NANO6 and h2ox1 with LeongRISK29 (PLAGE). Assuming that false positive non-progressors were due to random chance and that false positives would therefore be different individuals for different signatures, whereas the progressors would be positive across most signatures, requiring positivity on two signatures should decrease the number of false positives without unduly hampering the sensitivity.

GSVA scoring of h2otg+NANO6 had a PPV of 0.24 with 91% sensitivity, 88% specificity, and 31 false positives, compared to 50 false positives with h2otg by GSVA alone (Figure 4A). The combination of h2otg+NANO6 (GSVA) reduced the population-to-treat by 89% from 272 to 41. This combination was the best performing combination biomarker with only 12% of non-progressors being miscategorized, and achieved optimal WHO TPP for sensitivity. Similarly, h2otg+RISK11 in GSVA scoring had 100% sensitivity and 84% specificity with a PPV of 0.20 with 43 false positives. Combinations with h2ox1 also performed well under PLAGE scoring, with h2ox1+NANO6 achieving a PPV of 0.20, 82% sensitivity, and a specificity of 87% (Figure 4B). This combination had lower sensitivity and 2 missed progressors compared to the GSVA combinations with 0 or 1 missed progressors. Similarly, h2ox1+LeongRISK29 had a PPV of 0.18, a sensitivity of 82%, and a specificity of 84% (Figure 4B). This combination had 35 false positives and 2 missed progressors, compared to 46 false positives and 1 missed progressor with h2ox1 alone (Figure 4B).

**Figure 4:**
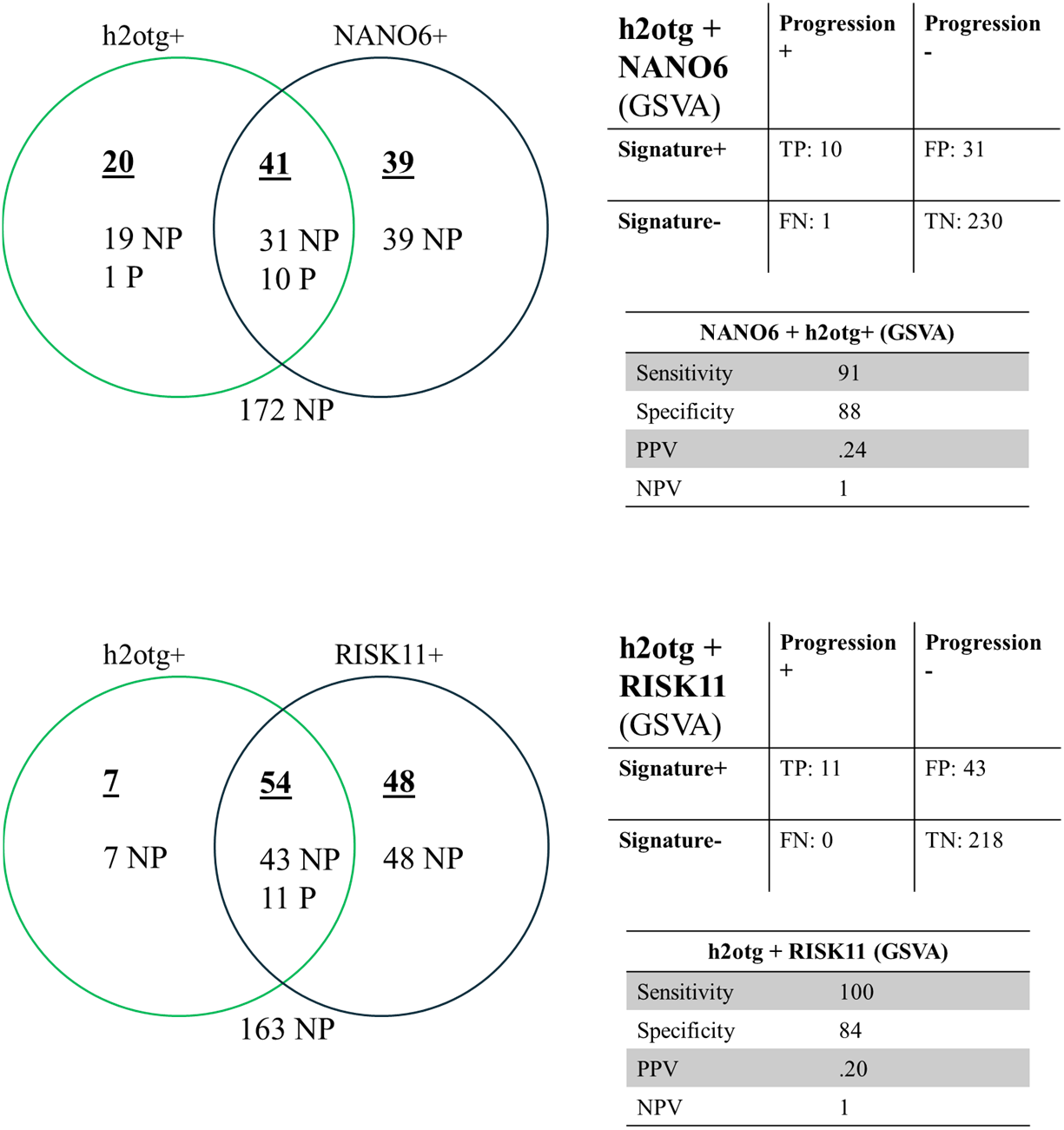
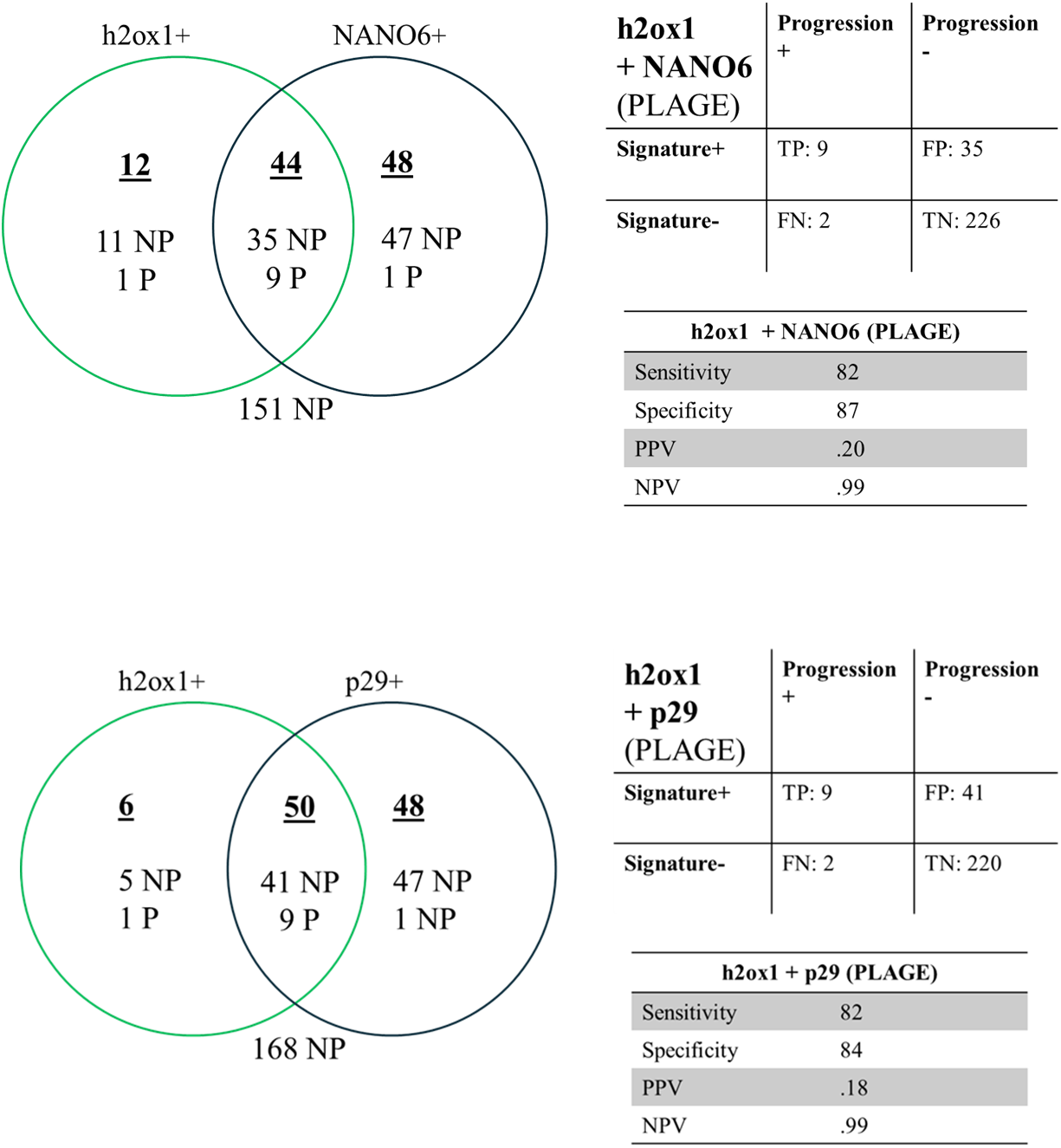
Combining Progression Signatures Increases Specificity. Venn diagram of HHCs positive for signatures, with progressors (P) and non-progressors (NP), confusion table, and performance metrics for signature combinations.

## DISCUSSION

One third of the world’s population is estimated to be latently infected with TB, of which 5-10% will progress to active disease within 2 years [23]. Biomarkers are urgently needed to identify those most at risk for targeted prophylaxis [24].

Several signatures have previously been identified that could predict progression to active disease in individuals with LTBI, including well-studied risk signatures RISK16, derived from the Adolescent Cohort Study (ACS) in South Africa, and Sweeney3, derived from multicohort analysis [1, 3]. However, neither of these signatures could accurately predict progression more than a year before disease onset, limiting their utility and increasing the chance of detecting subclinical TB rather than truly predicting progression at baseline. PREDICT29, a risk signature derived by our group from the Adolescent Cohort Study, predicted progression up to 5 years before progression, with an AUC of 0.911 in a Brazilian cohort [7]. Recent head-to-head comparisons of risk signatures using RT-PCR in a large Brazilian household contact cohort with 2-year follow-up found that risk signatures only met WHO minimum performance at 9 months lead time, not meeting the requirement for a 2-year prognostic window [25]. In our Brazilian cohort, which included progressors with a lead time to TB of up to 4 years, we found that when thresholding for high sensitivity cutoffs, the individual published signatures did not meet the minimum WHO specificity. However, combinations of newly derived parsimonious signatures and published signatures increased the specificity to reach WHO TPP levels. The recent comparison study in the Brazilian cohort did not include PREDICT29 or NANO6 signatures, both of which were top performers in our cohort, and while their study had a larger sample size, they had a much smaller percentage of progressors, 1.3% compared to 4.2% in our Brazil cohort, and included progressors without microbiological confirmation [25], which could explain the superior predictive performance of the combined signatures in our Brazil cohort.

NANO6, which distinguishes TB from LTBI with high accuracy [8], was found in this study, using GSVA, to be the top-performing signature in predicting progression to TB disease. In contrast, using PLAGE, PREDICT29 had the highest performance of the published signatures that were tested. Although most of the signatures had fair accuracy by AUC, the PPV values were limited due to the low incidence of progression to TB. In our cohort of 272 exposed HHCs, 11 progressed to disease (4%), determined by passive follow-up of the Brazil national TB registry, which was on par with the progression rate seen in studies with active follow-up of LTBI cases [4]. Although this limited the PPVs and resulted in NPVs near 1, an advantage of the observational study is that it approximates the true progression rate that would be seen if the test were utilized clinically. In both scoring methodologies, AUC values remained below 0.85, likely due to an inherent non-specificity of host-directed signatures, consistent with the unsupervised clustering data, which shows non-progressors exhibiting a similar gene expression pattern to progressors. NANO6 had the highest PPV of the published signatures (0.13) in GSVA, with 70 false positives, and reduced the potential prophylaxis candidates from 262 to 80. While this is useful as a triage test, in order to improve specificity, machine learning was used to generate new signatures in the dataset. Some h2o-derived signatures performed significantly better than published signatures with a PPV of up to 0.18, showing that h2o-derived signatures could retain accuracy without their model algorithm. Even the best h2o signatures resulted in 46-50 false positives and a maximum AUC of 0.89 (h2otg, GSVA).

Of particular significance, this study provides the first demonstration that combining signatures may improve specificity and PPV without sacrificing sensitivity. We found that h2otg and NANO6 in GSVA, the top-performing h2o signature and a published signature, respectively, had a combined sensitivity of 91%, specificity of 88%, and a PPV of 0.24, well above the WHO TPP minimum of 75% sensitivity and 75% specificity. Requiring double positivity reduced the false positives to 31 without missing more cases, with only 12% of non-progressors misclassified. Utilizing this combination as a triage test for preventative treatment could reduce the LTBI population to a reasonable size for intervention, while including those at the highest risk of progressing to active TB disease. RISK11 did not perform well in distinguishing between prevalent TB and viral respiratory infections [5], but by combining it with an h2o signature, it resulted in increased overall specificity, confirming the utility of combination gene signatures in triage tests.

Host-derived signatures are non-specific compared to microbiological tests. In our study, we observed that several non-progressors were double-positive for TB signatures with distinct genes, suggesting that these assumed false-positives were perhaps not due to chance. Possibilities include other lung pathologies or immune-perturbing conditions that weren’t captured at screening, or that those individuals will progress outside of the follow-up period. It is also possible that those individuals went on to subclinical disease that was self-cured or not detected and reported to the national registry. Because of the observational nature and passive following, it was not possible to capture subclinical disease to determine if signatures could predict progression to that state, a limitation of the study. Mathematical modelling of historical data has revealed that many individuals with LTBI develop subclinical disease without full progression to active TB [26].

Lastly, it is possible that those individuals mounted effective immune responses that cleared their infection, resulting in increased signal at baseline but preventing future progression.

Although the gene signatures are not highly specific compared to tests that detect Mtb, such as GeneXpert, the high sensitivity positions these tests to be clinically useful triage tests. Despite the concerns of side effects, non-adherence, and re-infection, some countries are pursuing aggressive LTBI treatment policies in response to the WHO call to end TB by 2030. By using h2otg+NANO6 as a triage test, which in our cohort reduced the HHC population to be treated by 89%, the population to receive prophylaxis could be greatly reduced while still ensuring most progressors receive treatment. This has direct clinical potential to reduce the incidence of isoniazid prophylaxis toxicity, as well as the potential for developing drug-resistant TB from patient non-adherence. Furthermore, by targeting treatment to those most likely to progress, this method can substantially reduce the costs associated with unnecessary therapy for non-progressors, particularly in resource-limited scenarios where healthcare budgets are constrained.

While we found that combinations of two signatures improve performance by increasing specificity, the larger number of genes to be profiled may be a limiting factor in clinical applications. In addition, while the NanoString platform and GSVA/PLAGE scoring are advantageous due to the unbiased approach and ability to multiplex TB signatures, they are not easily translatable to clinic due to requiring specific instrumentation. Smaller signatures have greater utility for point-of-care testing as they can fit on cartridge-based systems, such as Sweeney3, which was adapted to the Host Response Cepheid cartridge; latest versions of the cartridge can detect 10 genes [11]. However, despite multicohort validation, the small signature did not reach WHO target product profile (TPP) levels for prognosis at greater than 6 months in further studies [7, 27]. Larger gene signatures may potentially increase performance for predicting risk of disease progression by improving discrimination between TB disease progression and infection with other respiratory viruses/bacteria at the time of blood sampling. The latest versions of the Cepheid host response cartridge can detect 10 genes [11] and a 45-gene host gene expression test was developed using the BioFire System [28, 29], suggesting that in the future, these platforms could be utilized to develop rapid blood-based TB triage tests containing large gene signatures.

In conclusion, in this study, we used machine learning to derive a new parsimonious gene signature, which in combination with a published signature, showed increased specificity, meeting minimum WHO target product profiles, multiple years prior to TB onset. Upon validation, this combination test has potential clinical utility for identifying high-risk individuals for targeted prophylaxis to reduce morbidity and mortality and prevent the cycle of Mtb transmission.

## Data Availability

All data produced in the present study are available upon reasonable request to the authors

## AUTHOR CONTRIBUTIONS

S.L., R.R.R. and P.S. conceptualized the study; S.L. and V.K. contributed to data acquisition S.L. performed data analysis; S.L., R.R.R., W.E.J, J.J.E. and P.S. contributed to data interpretation; P.S., J.J.E., R.R.R., and R.D. acquired the funding for the study; S.L. and P.S. wrote the original draft; All authors reviewed and approved the final manuscript.

## Acknowledgements

This work was funded by the National Institute of Allergy and Infectious Diseases, National NJ ACTS Fellowship program Institutes of Health grants U19AI111276 to R.R.R., R.D., J.J.E. and P.S; S.L. received support from the NJ ACTS Fellowship program TL1TR003019. The study sponsors were not involved in the study design, in the collection, analysis, and interpretation of data; in the writing of the manuscript; or in the decision to submit the manuscript for publication.

