## Supplemental Data for "Combining blood transcriptomic signatures improves the prediction of progression to tuberculosis among household contacts in Brazil"

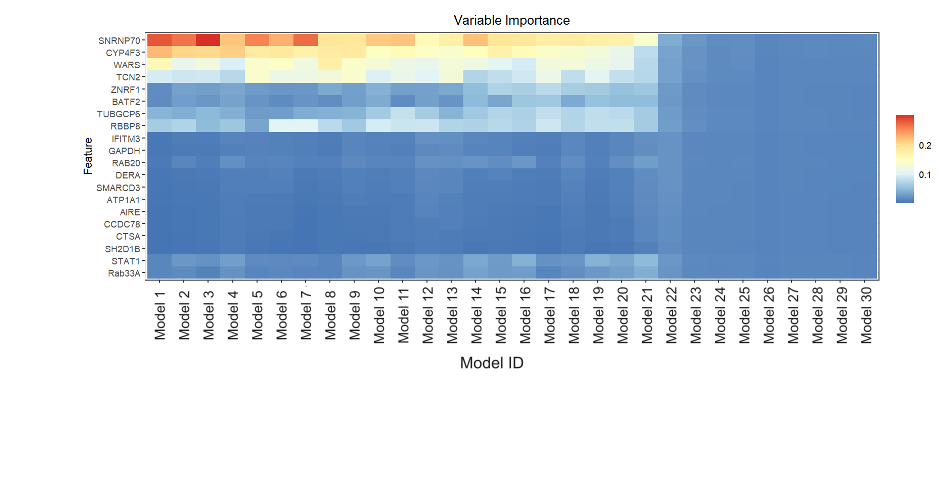

**Supplementary Figure 1**

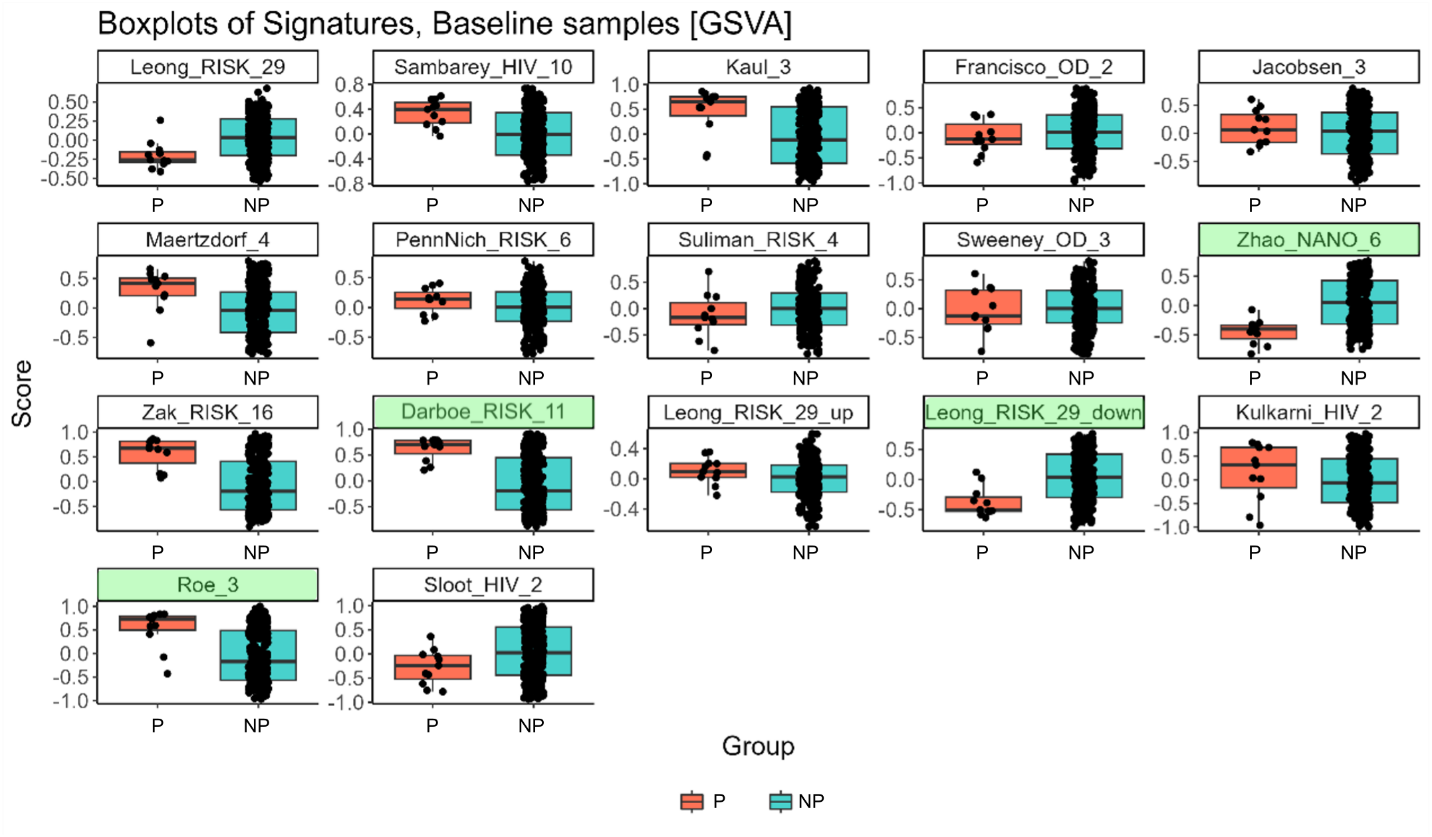

**Supplementary Figure 2**

**Supplementary Figure 3**

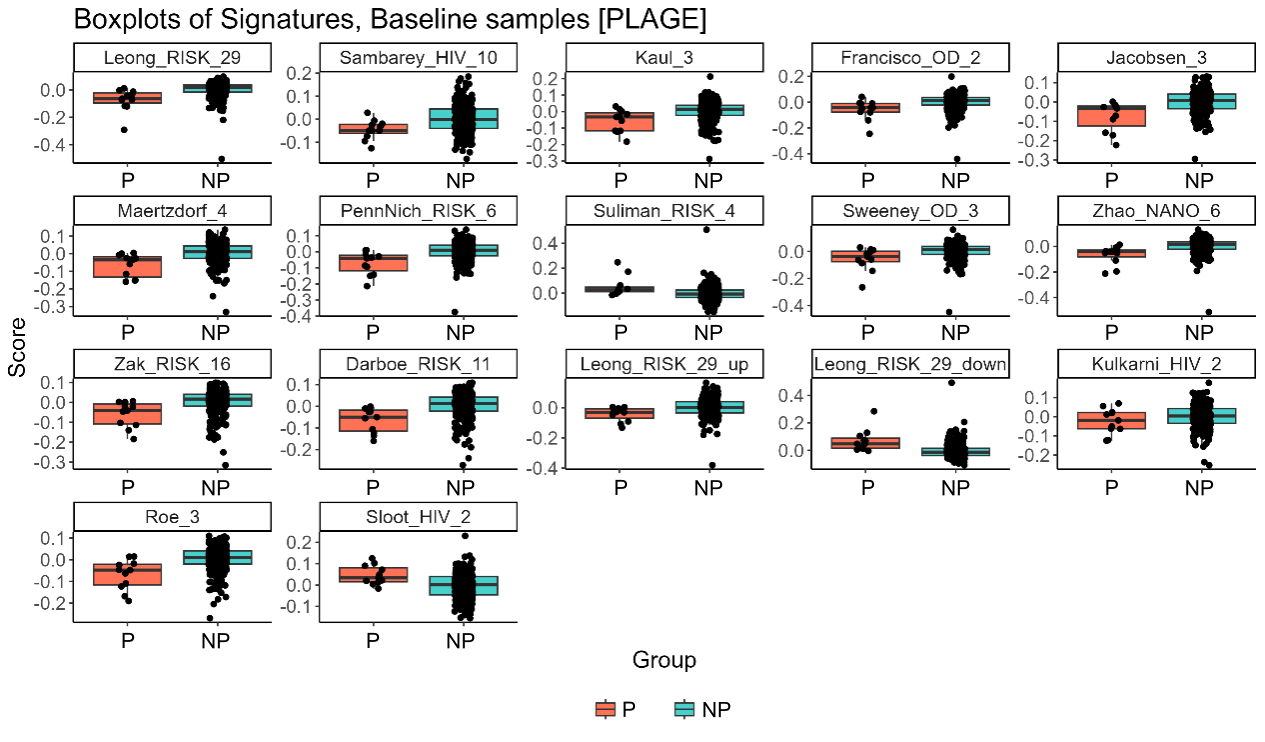

**Supplementary Table 1**: Participant Demographics.

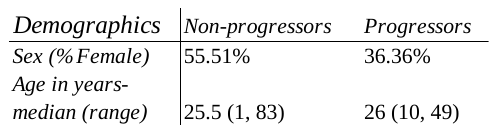

**Supplementary Table 2**: TB Disease Demographics

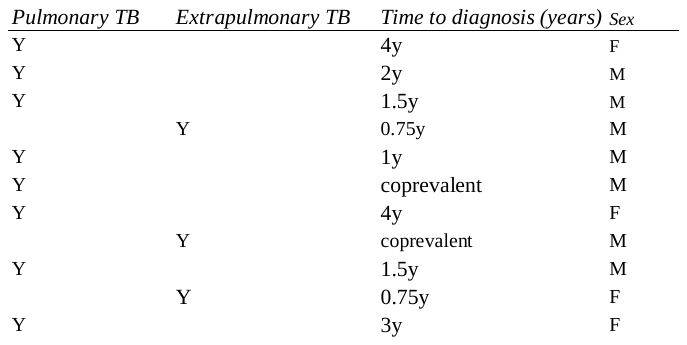

**Supplementary Table 3**: NanoString Codeset Signatures.

| **Citation** | **Signature** | **Comparison** | **Genes in Codeset** |
| --- | --- | --- | --- |
| Jacobsen et al | Jacobsen3 | TB/LTBI | CD64, LTF, RAB33A |
| Maertzdorf et al | Maertzdorf4 | TB/LTBI | GBP1, IFITM3, P2RY14, ID3 |
| Sweeney et al | Sweeney3 | TB/LTBI | DUSP3, GBP5, KLF2 |
| Sambarey et al | Sambarey10 | TB/LTBI | BCL6, CYP4F3, FCGR1A, HK3, IFI44L, RAB13, RBBP8, SLP1, SMARCD3, TIMM10 |
| Sloot et al | Sloot2 | Risk of progression to TB | AIRE, IL-13 |
| Zak et al | ACS-COR (RISK16) | Risk of progression to TB | ANKRD22, APOL1, BATF2, ETV7, FCGR1A, FCGR1B, GBP1, GBP2, GBP4, GBP5 SCARF1, SEPT4, SERPING1, STAT1, TAP1, TRAFD1 |
| Darboe et al | RISK11 | Risk of progression to TB | BATF2, ETV7, FCGR1B, GBP1, GBP2, GBP5, SCARF1, SERPING1, STAT1, TAP1, TRAFD1 |
| Suliman et al | RISK4 | Risk of progression to TB | BLK, CD1C, GAS6, SEPT4 |
| Leong et al | PREDICT29/RISK29 | Risk of progression to TB | AGAP9, APOL6, CCDC14, CCDC78. CIRBP, CTSA, DERA, ENO3, FUT4, GSTA4, HM14, IL31RA, LUC7L, MDN1, MOB3C, NAGA, NEIL1, PRSS63, SH2D1B, SLC3A1, SNRNP70, SPDYE5, SPSB1, SRBD1, TCN2, WARS, ZNF202, ZNF419, ZNRF1 |
| Kaipilyawar et al | NANO6 | TB/LTBI | ANKRD22, BLK, CCR6, ATP1A1, DARS3, EXOC2 |
| Kulkarni et al | Kulkarni2 | TB/no TB | RAB20, INSL3 |
| Francisco et al | Francisco2 | TB/other diseases | GBP5, KLF2 |
| Kaul et al | Kaul3 | LTBI/no TB | FCGR1B, GBP1, GBP5 |
| Penn-Nicholson et al | RISK6 | Risk of progression to TB, TB/LTBI | GBP2, FCGR1B, SERPING1, TUBGCP6, TRMT2A, SDR39U1 |
| Roe et al | Roe3 | Risk of progression to TB | BATF2, GBP5, SCARF1 |

**Supplementary Table 4**: Curated h2o-derived signatures

| **Signature** | **Genes** | **Curation** |
| --- | --- | --- |
| h2omlb | SNRNP70, CYP4F3, WARS, TCN2 | Top genes across 80% of leaderboard 1 models |
| h2omlb2 | SNRNP70, CYP4F3, WARS, TCN2, RBBP8, ZNRF1 | Top genes across 70% of leaderboard 1 models |
| h2oxlb | CYP4F3, ZNRF1, WARS | Top genes across 80% of leaderboard 2 models |
| h2oxlb2 | CYP4F3, ZNRF1, WARS, SNRNP70, RBBP8 | Top genes across 70% of leaderboard 2 models |
| h2oylb2 | SNRNP70, CYP4F3, ZNRF1, TCN2, RBBP8, HM13 | Top genes across 70% of leaderboard 3 models |
| h2ox1 | CYP4F3, ZNRF1, SNRNP70, TCN2 | Top genes (scaled importance>0.5) from top training model r2 |
| h2oy1 | SNRNP70, CYP4F3, ZNRF1 | Top genes (scaled importance>0.5) from top training model r3 |
| h2ot | TRAFD1, SMARCD3, GBP1, HK3, IFITM3, FCGR1A, CYP4F3, BATF2, GBP4, IL-13 | Top genes from best test performing model r2, without GAPDH |
| h2otg | TRAFD1, SMARCD3, GBP1, HK3, IFITM3, FCGR1A, CYP4F3, BATF2, GBP4, IL-13, GAPDH | h2ot with GAPDH |
| h2ot2 | ZNRF1, SMARCD3, IL-13, TRAFD1, STAT1, SRBD1, GBP4, SEPT4., HM13, AGAP9 | Top genes from best test performing model r3 |
| h2ompan | SNRNP70, CYP4F3, WARS, TCN2, STAT1, RAB13, AGAP9, GBP4, Rab33A, ETV7 | Top genes from different model types leaderboard 1 |
| h2oxpan | CYP4F3, ZNRF1, SNRNP70, WARS, SLPI, IL-13, IL31RA | Top genes from different model types, leaderboard 2 |
| h2oypan | SNRNP70, CYP4F3, ZNRF1, ETV7, SLPI, NAGA, TCN2, HM13 | Top genes from different model types leaderboard 3 |
| h2otestmnlbshort | BLK, CCDC78 | Best h2o-derived signature from a pediatric cohort, lacking one gene not in this codeset |

**Supplementary Table 5:** Ranked table of signature p-values and AUCs

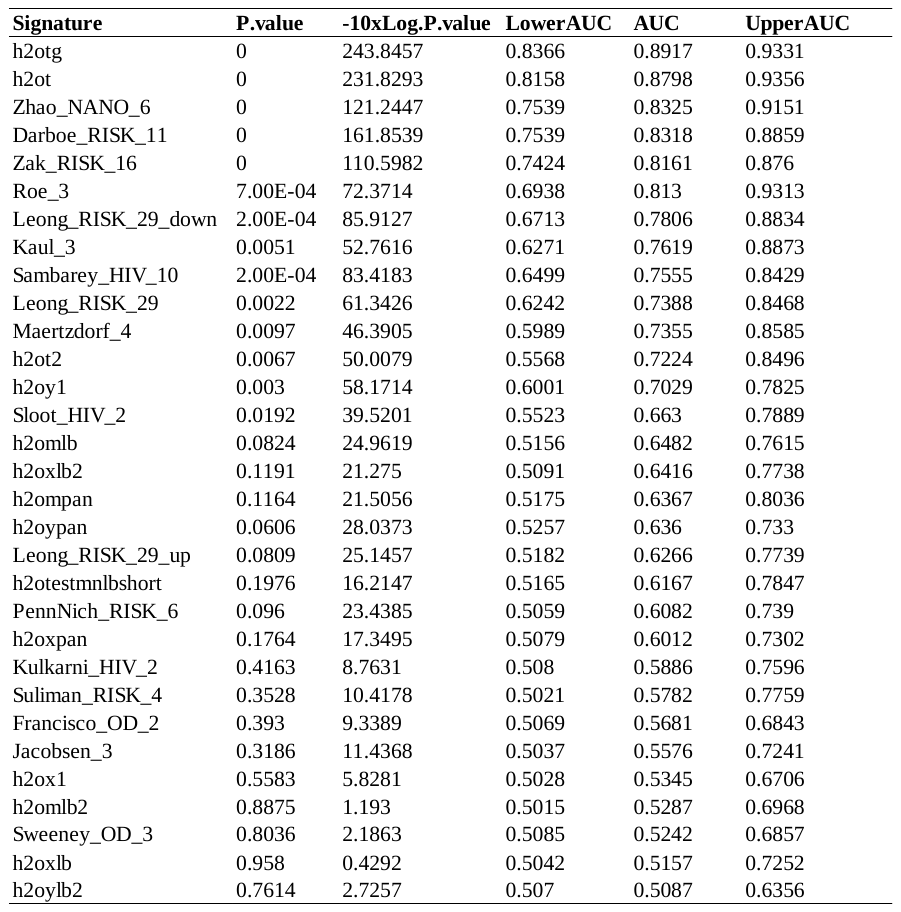

**Supplementary Table 6**: Ranked table of signature p-values and AUCs

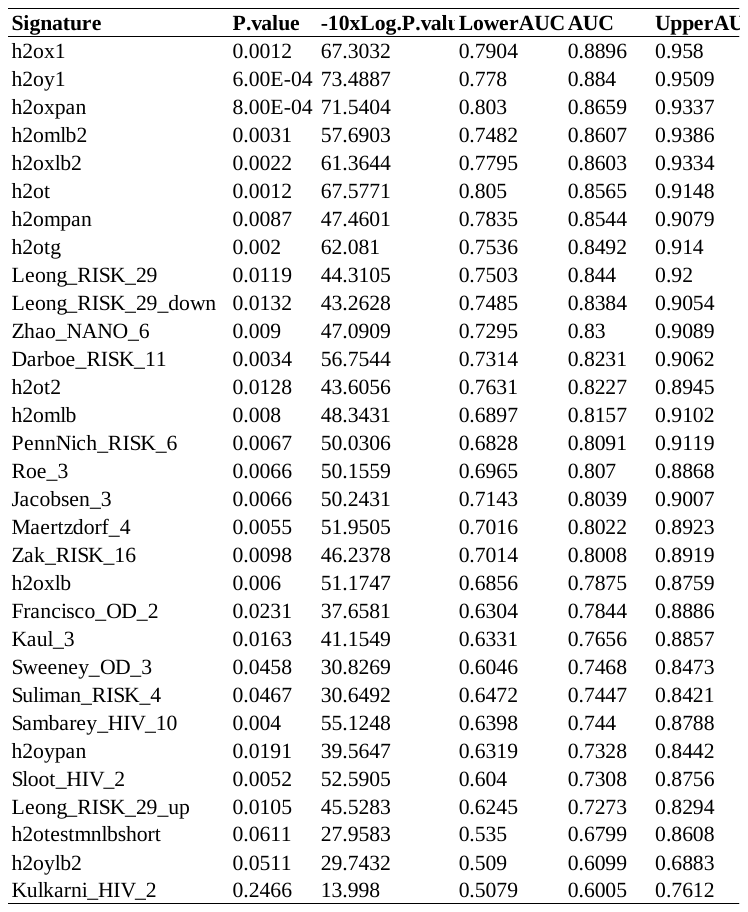
